# A roadmap for patient-public involvement and engagement (PPIE): Recounting the untold stories of breast cancer patient experiences

**DOI:** 10.1101/2023.06.19.23291192

**Authors:** Margaret R Cunningham, Nicholas JW Rattray, Yvonne McFadden, Domenica Berardi, Karim Daramy, Patricia E Kelly, Allison Galbraith, Isobel Lochiel, Lorraine Mills, Yvonne Scott, Susan Chalmers, Alison Lannigan, Zahra Rattray

## Abstract

**Introduction:** Breast cancer remains a prevalent disease in women worldwide. Though significant advancements in the standard of care for breast cancer have contributed to improved patient survival and quality of life, a breast cancer diagnosis and subsequent treatment interventions have a long-lasting impact on patients’ lived experiences. A high-quality healthcare system uses a patient-centred approach to healthcare, with patient engagement being a central pillar in the delivery of patient-centred care. However, the disconnect between patients and researchers can translate into research lacking real-world relevance to patient health needs. Here, we report a patient and stakeholder engagement workshop series that was conceptualized with the goal of promoting dialogue between patients with breast cancer, breast cancer researchers and the clinician involved in their care. We present the collaborative learning process and emerging opportunities from this patient engagement workshop series as a community-academic partnership.

**Method:** We report on a three-part storytelling workshop, with the scope of the workshops including topics related to raising awareness of the patient lived experience following a breast cancer diagnosis, breast cancer research activities undertaken by researchers, and the approach used by multidisciplinary healthcare teams in the management of breast cancer using storytelling as a tool. We used an iterative approach to cohort trust and relationship building, narrative development, and the use of multiple media formats to capture patient stories. This included the use of object memories, storytelling prompt cards and open-mic audio format to capture patient stories from diagnosis to treatment, and remission.

**Results:** 20 patients shared their stories with key themes emerging from the qualitative analysis of audio recordings. For many, this was the first time they had spoken about their breast cancer experience beyond family and friends. Emerging themes included common public misconceptions about a breast cancer diagnosis, the importance of self-advocacy in patient decision making about treatment, and the complex emotional journey experienced by patients diagnosed with breast cancer. The group-based storytelling approach provided collective empowerment to share personal experiences and connect meaningfully across the peer community.

**Conclusion:** While a breast cancer diagnosis can be overwhelming from a physical, social, emotional and cognitive perspective, storytelling as a patient engagement approach can build patient trust in researchers, ensuring that as key stakeholders they are involved in the process of research. Understanding the patient perspective of a breast cancer diagnosis and subsequent experiences can support healthcare professionals in developing an empathetic approach to sharing information, and involving patients in shared decision making about their healthcare.

## Introduction

Breast cancer remains a leading cause of female cancer-related diagnoses in the UK, with up to 4,200 new cases diagnosed in Scotland per annum (PHS 2021). Advancements in early screening campaigns (mammography), standard of care interventions, and patient education has seen significant improvements in patient prognosis beyond a breast cancer diagnosis (Ghoncheh, Pournamdar et al. 2016, Wojtyla, Bertuccio et al. 2021). The term ‘breast cancer’ represents a heterogenous collection of diseases, with breast cancer classification occurring based on molecular subphenotypes, hormone receptor expression, and histological presentation, each of which dictate the use of different intervention pathways for patients. These intervention pathways will often require the use of multiple therapeutic interventions such as surgery, chemotherapy, radiotherapy, or the use of endocrine therapies, each of which are associated with different long-term effects that may impact patient quality of life years beyond their care (Fraser 2022).

While recent technological advancements have revolutionized the diagnosis and treatment of breast cancer, these advances alone cannot be relied on to support a patient in their journey of illness. Patients become entangled in the confusion of scientific jargon, the fear of unknown, and lack of effective communication with healthcare professionals about their perceptions, emotions, and lived experiences (Hoddinott, Pollock et al. 2018). Models of healthcare have evolved from a paternalistic model to patient-centred approach, where healthcare professionals are encouraged to implement a holistic approach. Despite the widespread implementation of patient-centred healthcare practices, research suggests ethical erosion among practitioners, with a decline in empathy and sympathy with increasing clinical experience (Liao and Wang 2020). Increased pressure on healthcare systems has created an environment resulting in less-exhaustive clinical examinations, focusing on symptom management where the provision of lifestyle advice is adversely impacted (Tsiga, Panagopoulou et al. 2013).

Clinical care leads to the co-creation of narrative that is formed and transformed by the encounter between the patient and healthcare professional, which evolves during the different stages of their illness. Healthcare professionals are trained in the art of history taking to extract relevant information efficiently, without spending time on unnecessary detail. However, permitting narrative flow during the consultation does not use substantial clinician time. Previous work has shown that two minutes is sufficient time for 80% of patients to voice their concerns, with ∼2% only requiring more than five minutes (Kalitzkus and Matthiessen 2009).

In recent years patient and public involvement and engagement have come into focus from funding bodies and healthcare-based research institutions to drive stakeholder driven input into research prioritization and initiatives (Hoddinott, Pollock et al. 2018). At the time of writing this manuscript, many UK charity healthcare research funding bodies (e.g., Breast Cancer NOW, Dunhill Medical Trust), require the inclusion of a patient and public involvement (PPI) statement and plan in proposal submissions as primary assessment criteria. The value of patient and public involvement and engagement (PPIE) is becoming widely recognized as an integral part of research, not only at the mid-end stage of project development, but as essential stakeholders at the point of project conception and design. Most of these efforts are normally focused on specific and individual research projects or programmes receiving input and feedback on their proposed vision and activities from patient experts or advocates. While PPI has proven benefits in the inclusion of the patient voice in research, it can be heavily influenced by the research team narrative or those patients with extensive prior involvement with researchers (Winterbottom, Bekker et al. 2008). Public and patient engagement provides researchers with the opportunity to share their research with citizens and enable a two-way dialogue between the audiences and researchers. This model allows for a flexible approach to patient involvement, enabling both feedback on current efforts and inspiration for future research priorities (Novak, George et al. 2020). Though extensive guidelines and training resources exist on how to conduct PPIE, there are a paucity of publications or case studies providing exemplars of such activities and to a lesser extent the approach that was used to conduct PPIE.

Storytelling has become increasingly used a pedagogical tool to effectively engage the public with science (JE, S et al. 2022), however this can often be poorly designed in a way that leads to one-way communication and lack participatory input. Therefore, the design of activities that promote interaction, reflection and authentic story development within a patient cohort setting is critical. The approaches used in the current study to promote engagement included object-based storytelling using mementos brought by each participant. This is a common storytelling approach used to foster positive conversation between people during group activities through displaying visual mementos which are later used as focal points for discussion (Winterbottom, Bekker et al. 2008). Narratives were then visualised to promote reflective practice (Narelle 2006) and provide source material and evoke memories from which stories could develop and be reconstructed for oral communication.

Here, we present the collaborative learning process, the approach, and key findings from a workshop series in a breast cancer survivor storytelling project. The aims of this project were to break down the barriers between researchers, patients, and clinicians, facilitating the development of new relationships, while simultaneously learning about patient health concerns and perceptions of breast cancer. We used patient stories to develop a list of research questions, as well as considerations for patient counselling and education following a breast cancer diagnosis. To achieve the aims of this process, we developed an environment in which patients can rationalize their lived experiences of breast cancer with peers.

### Structure of the event

#### Recruitment

All participants in the three-stage storytelling workshop were women diagnosed with breast cancer after 2020 and treated in NHS Lanarkshire. The potential participants for the workshop were identified by the breast cancer consultant involved in their care. They were issued an invitation letter by the breast cancer surgeon to participate in a three stage storytelling workshop series, which included a participant information leaflet in which patients were informed of the process involved during the workshops and an associated written consent form to distribute media generated from the workshop. Those who agreed to receive further information regarding the workshop series or participate in the workshop series, provided their contact details in the form of phone numbers, email addresses and postal addresses.

#### Participants

A total of >30 patients were invited to collaborate and 20 agreed to participate, with seven scientist participants (various career stages) and the lead breast cancer clinician. Of the participants, eight patients attended all workshop sessions based on availability. All participant quotes, feedback, audio recordings and artwork were anonymized to protect patient identity.

#### Workshop format

Starting in August 2022, a three-part storytelling workshop was conducted with patient participants and researchers, with the breast cancer surgeon being the connect between both with prior engagement with patient and research participants. A workflow of the workshop format is shown in Figure 2.

**Figure 1.**
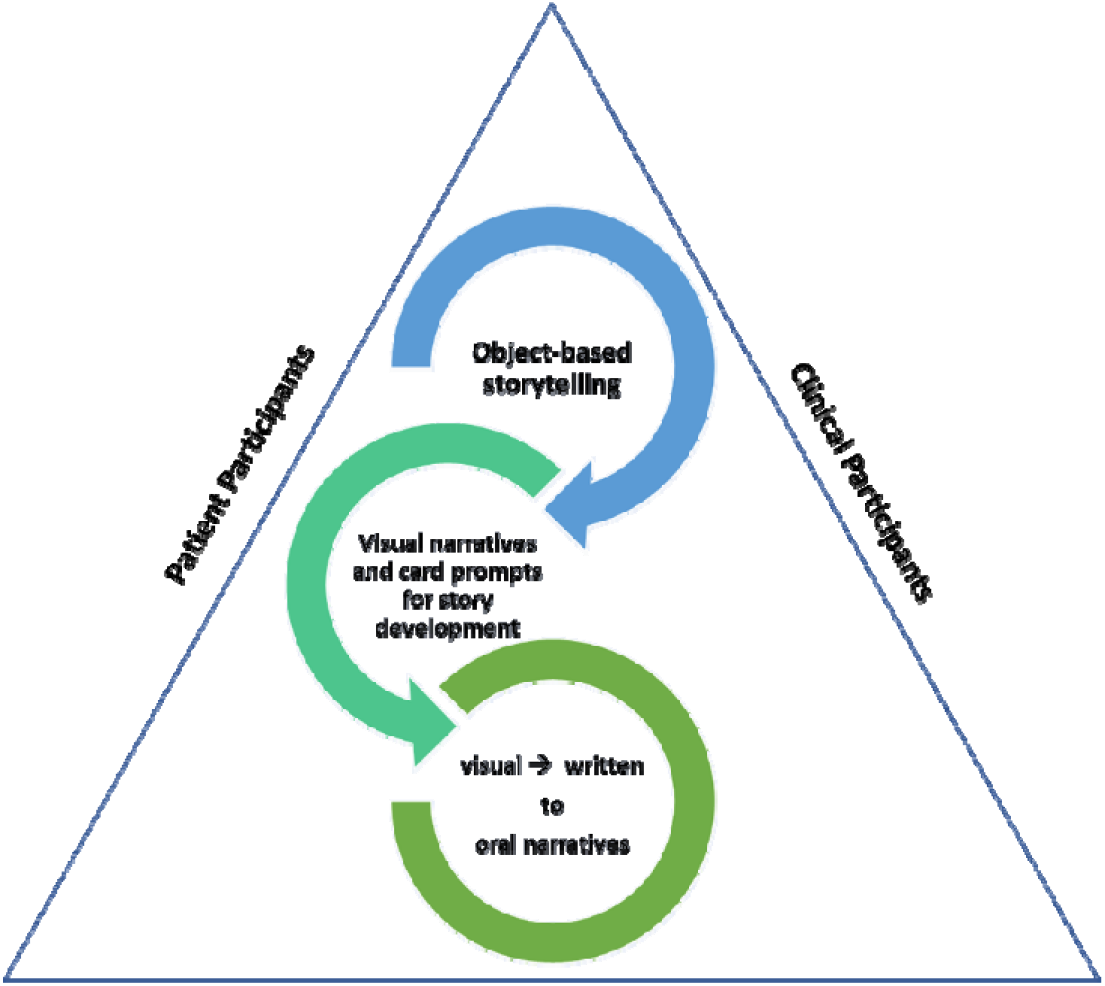
Using storytelling tools to establish meaningful communication across patient, clinical and scientist workshop participants.

**Figure 2.**
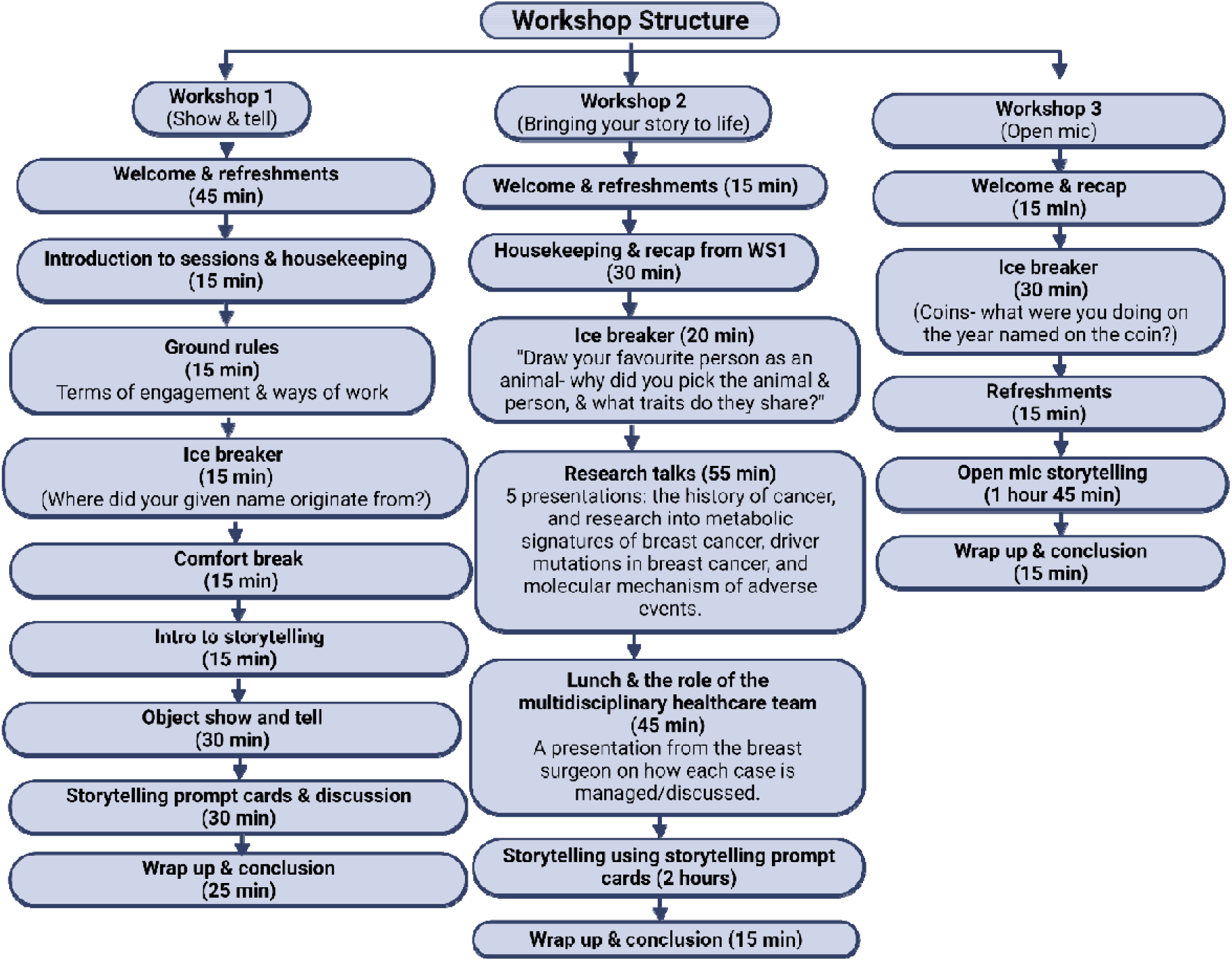
Workshop schedule and format used to build trust and participant narratives.

### Workshop 1- “Show and tell”

The aim of workshop 1 was for the patient and scientist participants to meet each other and develop a cohort. Feedback from participants on the first session, was used to inform the design of the second workshop.

During workshop 1 all participants were encouraged to network with their peers and scientists. This was followed by defining the ground rules for terms of engagement between participants. All participants reflected on instances of past experiences, and what constitutes a safe space and positive environment when discussing potentially emotive subjects (i.e., lived experiences). Following interactive ground rule setting, a brief interval of training on storytelling and the stages involved in the development of narratives was covered to introduce participants to storytelling and narrative development. To enable the use of effective storytelling practices, we partnered with a storytelling professional (Allison Galbraith from the Scottish Storytelling Centre) to help participants identify, share, and structure stories of their personal lived experiences in a personalized and effective way.

All participants (patients, researchers, clinician) were prompted in advance of the workshop by email communication to bring an object of personal significance, which would be used to capture the role of object memory in the lived experience and shared their stories within small focus groups of 5-7 participants. For those who did not have any objects, an object table (e.g., books, crochet, bell) was provided to ensure inclusion of all participants in this activity.

In response to the prompt to bring objects of significance, all participants brought an object and in instances where they had no objects available, a table of objects was provided to enable inclusion of all participants in object-aided storytelling.

Stories captured from object memory discussions shared by patient participants were captured and converted to visuals (Figure 5). Recurring objects selected from the object-aided storytelling were patient wigs, books that played a central role in coping during treatment, photographs and pictures recalling memories from their breast cancer journeys. Additional objects included a camisole to signify dignity and privacy during the tattooing process experienced during radiation therapy, and a bell to signify the end of cancer treatment.

**Figure 3.**
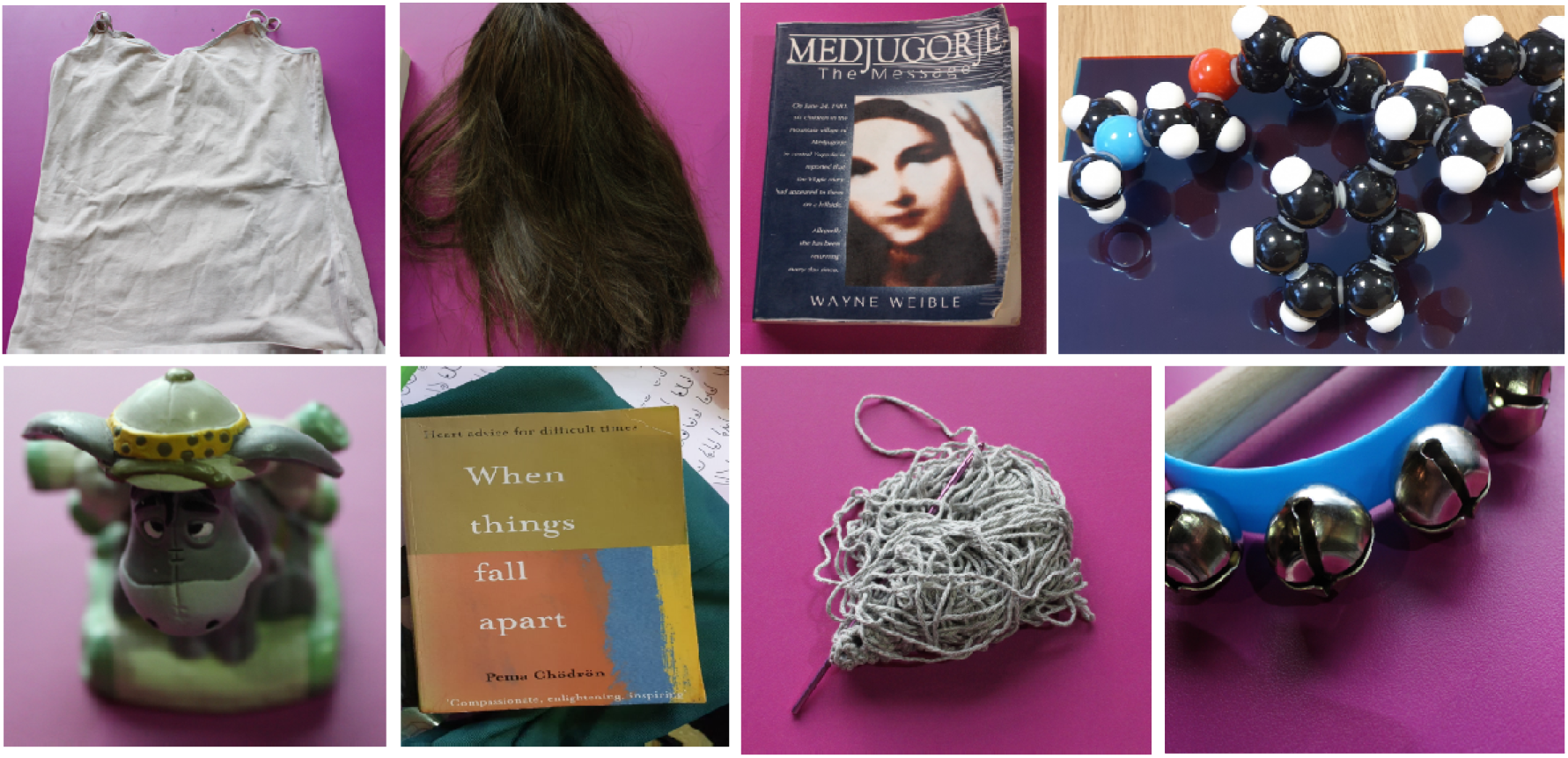
Exemplar objects that patient participants presented on their lived experiences.

**Figure 4.**
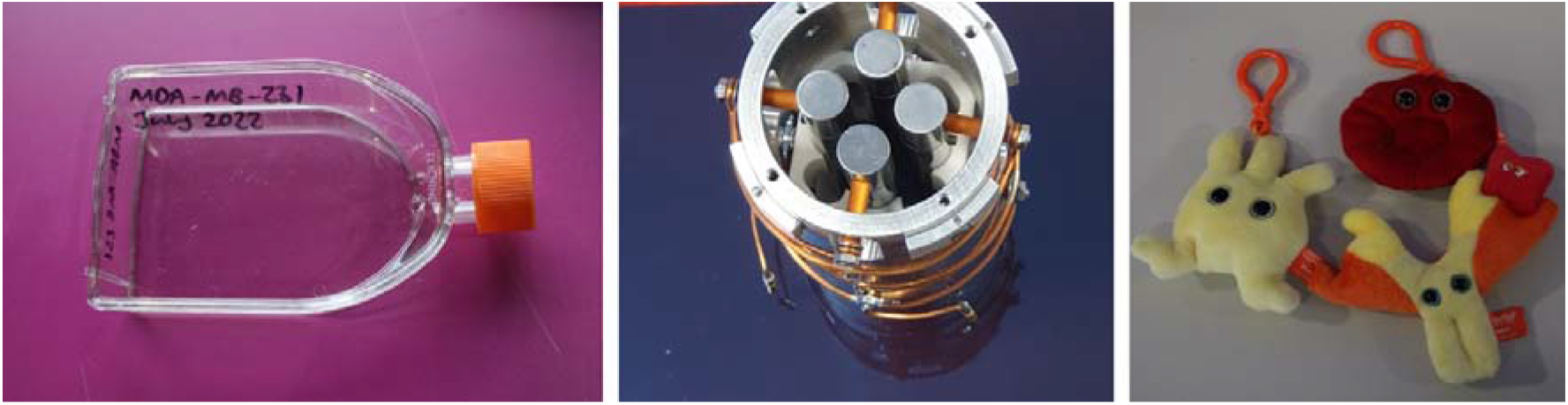
Exemplar objects that research participants presented on their work.

**Figure 5.**
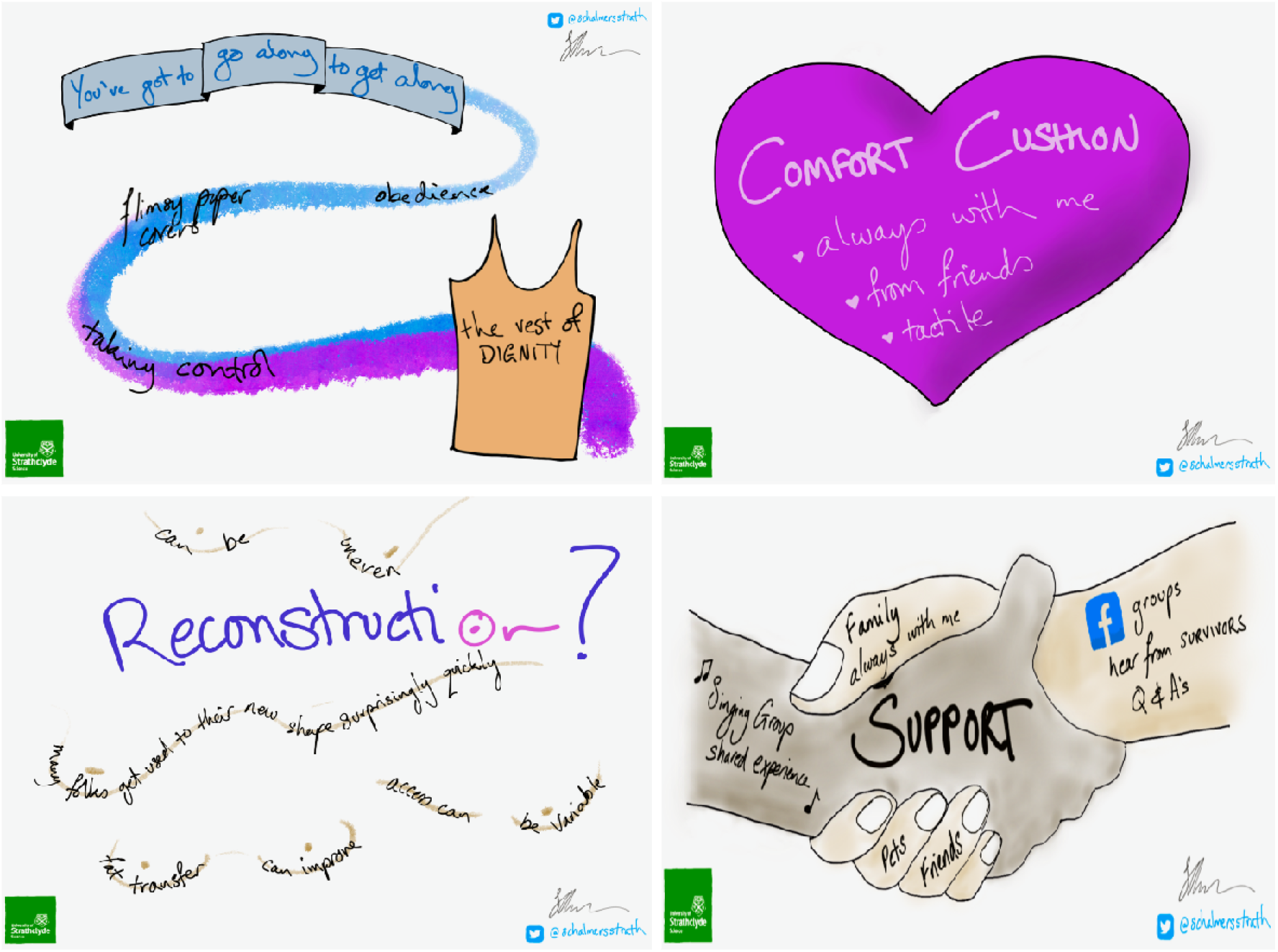
Exemplar visual narratives created from patient object memory stories

**Figure 6.**
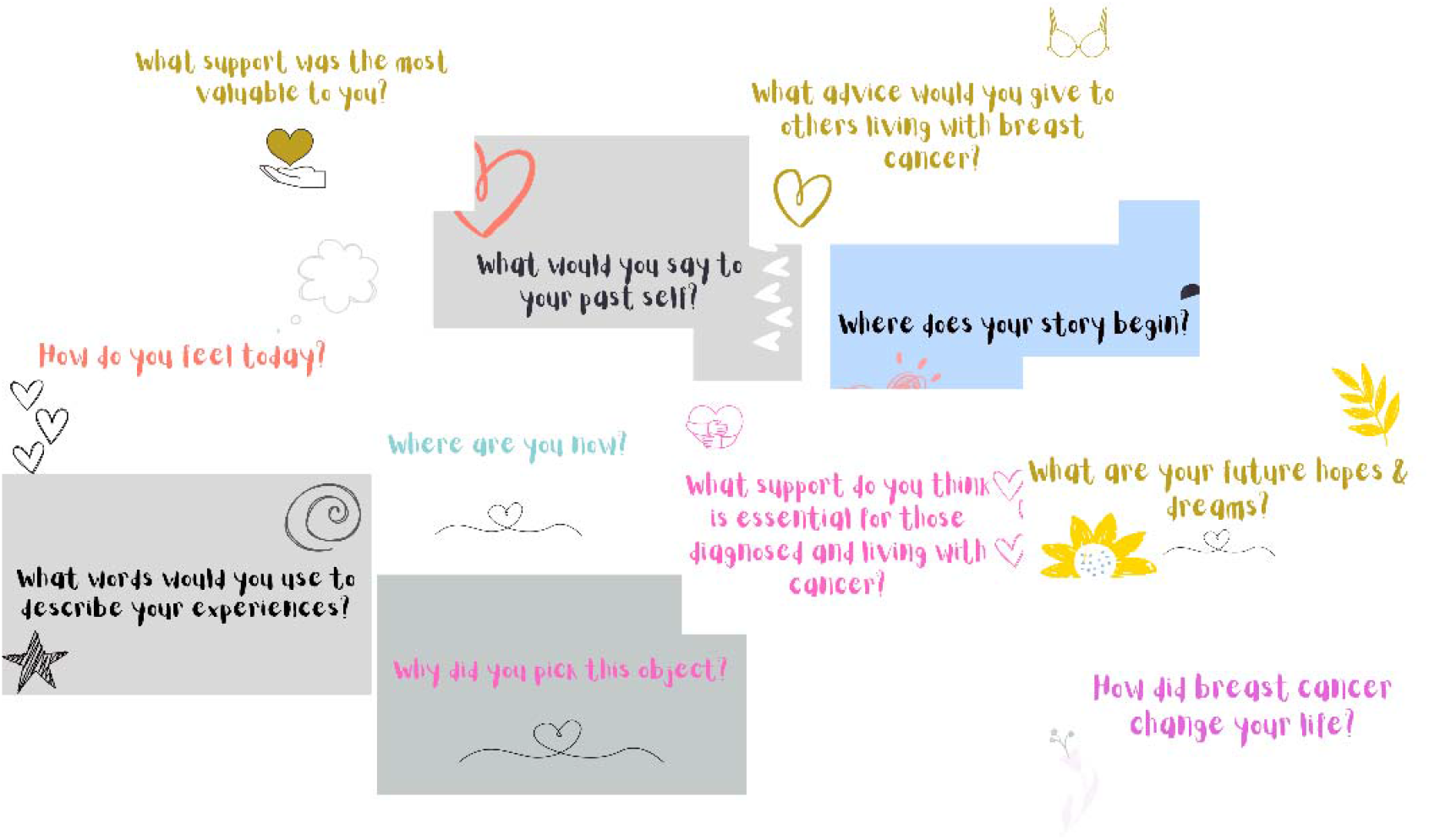
Prompt cards used to support patient participant narrative development during the workshop

The scientists presented objects that they use in their research (a tissue culture green lab coat, component of a mass spectrometer, and a cell culture flask labelled with a breast cancer cell line) and an ornament that the clinician presented to share a memorable patient encounter.

Following object-based narrative development within smaller focused groups, all participants placed their objects in a central table and shared their narrative with the rest of the participants. As everyone recalled their object memories, a member of the facilitator team was simultaneously visualizing participant stories and emerging themes. Visual narratives are a powerful way to promote reflective practice in group settings and develop confidence when interacting with peers Workshop 1 concluded with a reflection of all activities, followed by an introduction to activities for workshop 2, including timings and details.

A feedback notebook was provided for all participants to add their reflections and feedback on how the session went. Any feedback provided, was used to design subsequent workshop events.

### Workshop 2- “Bringing your story to life”

In workshop 2, breast cancer researchers and the clinician presented their work in story format, starting with the history of breast cancer, its diagnosis and treatment. This was followed by researcher facilitators sharing their research in story format covering various topics such as the role of driver mutations in breast cancer prognosis, mass spectrometry-based metabolomics of patient samples to fingerprint tumours, and studying the drivers of chemotherapy-induced cardiotoxicity as exemplars. These talks were designed to be interactive, where all participants could ask questions.

The research talks were followed by a demonstration from the clinician who described the role of multidisciplinary teams in healthcare decision making and the selection of treatment interventions. In the latter half of the workshop, the patients were given storytelling prompt cards to consider the beginning, middle and end of their breast cancer story.

Used within small breakout groups, the purpose of using storytelling prompt cards was to support the development of patient narratives and ensure equal and representative engagement from all participants (patients, researchers and clinician). Facilitators within breakout groups, were responsible for capturing timelines of patient stories and all breakout groups shared their relative experiences with all participants attending the workshop. During this session, participant stories were captured using multiple media formats (quotes, timelines, drawings, and an essay).

In the third workshop session, patient participants were invited to share their lived experiences of breast cancer in an open-mic format.

### Workshop 3- “Open-mic”

In the third and final session, there was an exhibition of photographs from all past workshops, including patient quotes, and imagery from their story arcs. All participants were invited to present their story in open mic format, while seated and the microphone was passed between participant volunteers throughout the session. All stories were captured in audio format with participant consent. The aim of this final session was for patients to voice their lived experiences, sharing their breast cancer diagnosis and treatment experiences with their peers. Following the completion of open-mic storytelling, key emerging themes and concepts from the patient stories were discussed within the group. The workshop concluded with feedback on the delivery of the workshop, and how it had benefited patient, clinician, and scientist participants in different ways.

### Logistical considerations for the design of a PPIE event

Patients invited for participation of this workshop were geographically based within the Lanarkshire region, which is situated outside the borders of Greater Glasgow. These communities are often underrepresented in University PPIE and outreach activities, due to the need for travel or challenges in encouraging participants to travel into the city to participate in such events. We also perceived that meeting the patient participants within a University campus could provide an additional barrier when they were already concerned about voicing and sharing their emotions. Accounting for these considerations, we decided to select a public venue within the local community for the workshops that would be accessible for all participants and provide an environment in which a safe space and place of trust can be created. Through our engagements with North Lanarkshire Council, we hosted all three workshops at the SummerLee Museum of Scottish Industrial Life. An advantage of selecting this venue was convenience in accessibility, the availability of appropriate facilities (accessible toilets, lifts), and readily accessible links to public transport.

## Discussion

Communication between patients, healthcare professionals, and researchers is increasingly being considered in cancer research, an area which has seen ever-increasing emphasis on PPIE in the design and dissemination of research activities to key stakeholders. To-date many participatory approaches have been implemented in PPI for cancer research, to enable communication between researchers and patients within the confines of a predefined project remit or subject area (Pii, Schou et al. 2019). This is in part a consequence of the rapid growth in clinical and translational research, and the need to ensure that technological advancements made in diagnostics and therapeutics are directly aligned with the needs of the target patient population. A major challenge that remains with existing approaches to PPI is the lack of critical reflection of process. Various models of PPIE have been proposed, that are defined based on the scope of patient involvement and can be broadly categorized as a ladder with the rungs of engagement ranging from informational (passive participation) to controlled (defining the direction and priorities in research) interactions depending on the nature of PPIE activity or the body involved in its design (Arnstein 1969). Reflective practice in the PPIE process was factored into the design of the approaches used in the current study in a way that each workshop was informed by the experiences captured in the preceding event.

PPIE activities may take place during various stages of research and concept design including research focus, research design, recruitment, data generation and analysis, and dissemination activities (Pii, Schou et al. 2019). Several systematic reviews have highlighted that PPI activities may take place across one or multiple stages of research project conceptualization and delivery, with most reviews of participation in cancer research indicating more PPIE occurs during initial stages of research design for defining research priorities. This often means that the impact of such activities is often lost through lack of patient engagement at the later stages of the research project. The long term loss of which can give the impression that PPIE activities are tokenistic in scope (Corner, Wright et al. 2007). In the current study, all participants were provided with equal opportunity to contribute towards projects emerging from the discussions, feedback and stories shared. So far this has also extended to the analysis of the qualitative data generated and provision of peer feedback on resources developed from material generated during the workshop by undergraduate students as part of their final year projects. The ethos of the team approach has been to nurture relationships across all stakeholders from co-creation to publication and form a PPIE working group for involvement in research proposals and education-based projects to enhance teaching and learning.

A key aspect that has been identified from systematic analysis of cancer PPI initiatives is the lack of reporting on challenges and recommendations, nor sharing of processes that were followed during the PPI activity. Therefore, the longer-term legacy and impact on subsequent PPI activities is often lost (Pii, Schou et al. 2019). Moreover, an element that remains unaddressed in many PPIE activities, is “how do we empower patients to have a real voice?” and “what is the best approach?”, to ensure that the patient voice is heard during PPIE initiatives, and that they also directly benefit from such activities in a more active capacity. As part of the workshop series findings, we report both the positive and negative impact of the experiences shared as part of the workshop series. Many of the patient participants cited that they had been in denial of having cancer and that they had not really visualised themselves as being a cancer survivor until the workshop experience. One participant even acknowledged that the realisation of their survivorship status had prompted them to seek additional support from their local cancer charity. This will be a perennial challenge with PPIE as it may evoke situations that may lead to patients re-living negative experiences or coming to terms with the illnesses that they have endured therefore post-workshop engagement and follow-up is essential.

The rise of widespread PPIE activities and emphasis on their implementation in research, has seen the development of a range of educational resources and toolkits to researchers, through public partnerships, UK-based charities and trusts (e.g., the Cancer Research UK patient involvement toolkit) (CRUK). While these resources are informative for engaging patients, they are often written by researchers and healthcare professionals with the patient in mind and do not provide a direct template for the successful design and delivery of a PPIE event. One of the primary developments that emerged immediately from the workshop experience was the design and development of education-based projects for the purposes of information resource design. These co-created projects are ongoing as part of the qualitative analysis carried out from material generated during the workshop series.

In the design of this storytelling workshop series, we aimed to overcome some of the limitations described above by engaging an audience who had not previously served as an expert patient, or an advocate. All participants were provided with the same training on storytelling. Storytelling as an aid, facilitated meaningful communication of participant lived experience narratives and successfully led to the step-wise development of rich stories of patient experiences.

Moreover, all patient participants in our workshops had been diagnosed during the course of the COVID-19 pandemic, where they had limited access to peer support networks or the opportunity to meet with other patients with breast cancer. Overall, the workshop provided the participants with the opportunity to connect not only with researchers, but those that had a similar lived experience throughout the course of their diagnosis and treatment for breast cancer.

Here we discuss some of the lessons learned from the design and delivery of such a PPIE event, where we have grouped these according to key concepts.

### Design of workshop series

#### The team

The concept of the workshop was initiated by the clinician and lead author(s). During initial scoping meetings, the requirement for additional skillsets (i.e., storytelling, oral history, and live sketching, and volunteer gender balance and intergenerational facilitators) was discussed to ensure that the appropriate team structure was developed to deliver on the workshop concept objectives.

#### Number and timing of workshops

Through engagement with the facilitator design group and patient participants, we decided to host the workshop series over three events to maximize the benefit to the patients and build trust and confidence. The timings of these events were designed to accommodate those with caring responsibilities, those in employment, and accounting for travel to the site, at low demand times (August-September). As a result, two 3.5-hour sessions (workshops 1and 3), and one 6-hour (workshop 2) session was developed.

#### Facilitators

There were several researcher participants involved in the facilitation and delivery of the workshop series to ensure representation and the streamlined delivery of workshop activities. For each of the workshop events, there were seven researcher facilitators (four academics and three doctoral students). The representation of the researcher community also served as a good opportunity to promote dialogue and networking with patient participants during the icebreaker and refreshment intervals. Overall, these interactions created a safe space and levels of trust and comfort throughout the cross-section of all participants attending the workshop series. Beyond the researcher participants being embedded within smaller group activities within their respective breakout groups and underpinning the delivery of activities smoothly, they would also alert the lead facilitator in the event of any issues arising (e.g., taking patients to the quiet safe space if recounting their experiences became too emotionally taxing).

#### Participant recruitment and cohort development

Two approaches were used to recruit patient participants for the workshop series. The first approach was to use an open invitation to participation *via* a poster promoted on various social media platforms (LinkedIn, Facebook, Twitter) with a QR code directing the reader to an Eventbrite webpage. The second approach was implemented *via* an invitation letter and participant information sheet issued to all breast cancer patients attending the NHS Lanarkshire breast clinic. All participants invited to the workshop series had been diagnosed and treated for their breast cancer in NHS Lanarkshire under the care of the same breast cancer surgeon. The latter approach was directly successful in translating to attendance on the workshop days. Overall, 20 patients participated across the three workshops, with eight attending all three workshops, 12 attending two workshops, and 10 attending the third workshop. Each workshop was designed to be sufficiently standalone, so that patient participants could recount their full story in workshops 1 and 2 if they were unable to attend the third workshop.

Another aspect that was considered during the workshop design stage, was to rotate the composition of the participant groups (patients, researchers, facilitators) across every activity and session, to promote extensive networking and overcome any potential issues arising from participants feeling uncomfortable in each other’s presence.

#### Design of workshop activities

Throughout the course of the workshop series, we implemented a combination of discussion-based and task-based activities. The discussions would take place following task-oriented activities in smaller breakout groups. We found that the sequential combination of task followed by discussion allowed for every participant to feel confident and comfortable in contributing to the wider group discussions. For example, patient participants were asked to start capturing the key timelines in their breast cancer stories as a task using storytelling prompt cards and were provided with transparent acetate sheets. Taking turns around the table, each participant would describe their sketch and timelines to their peers. All participants would share aspects of their stories that were common and relatable, while they would also describe the unexpected differences in their breast cancer experiences.

#### Feedback

All participants were provided with a booklet in which they could provide feedback on the event, or suggestions for the subsequent workshops. Feedback received from all participants indicated that though the initial thought of sharing their breast cancer lived experiences was daunting and concerning, the workshop series provided them with a positive safe space in which they could find others with similar experiences. They also agreed that they had developed a better understanding of the range of breast cancer research activities taking place and how these will contribute to the improved standard of care for patients with breast cancer in the future. Examples of direct feedback quotes provided at the end of the sessions from patients are included as follows:

> *“I feel that the learning from this workshop must be heard by other breast cancer patients who are starting their own journey. This would have been beneficial to me-would like less listening and more group work i.e. experiences.” Participant from workshop 1 (Aug 2*^*nd*^ *2022)*
>
> *“It was CATHARTIC. The format worked very well with our lovely group. A very human and compassionate experience. <3” Participant from workshop 1 (August 2nd 2022)*.
>
> *“When the MDT process was described at the workshop, it made me feel safe knowing that my case is being discussed in detail as a person. It was great to hear about the process” Participant from workshop 2 (August 18*^*th*^ *2022)*.
>
> *“I attended the event as the husband of a breast cancer survivor and would encourage other husbands/partners to attend future events. The day was educational interesting and inspiring, and I am pleased that I chose to attend. I loved learning other women’s stories and have a huge admiration for the bravery, stoicism, and humour that they demonstrated when discussing their own personal experiences.” Participant from workshop 2 (August 18*^*th*^ *2022)*

#### Legacy and limitations

Overall, the response from all participants was overwhelmingly positive, with most patients expressing that they did not wish for the workshop series to end, and that the event had a long-lasting positive impact on their mental health and wellbeing. A limitation of this workshop series was that participants who responded were all of Caucasian descent, with lack of representation from other minority ethnic groups who were also invited. The event series was very informative in identifying health education needs for patients presenting with a new breast cancer diagnosis and creating a community of peers. However, with the event series reaching a natural end, the patients felt isolated. Future work would consider inviting the participants to the University campus or support the development of patient groups through the community centres.

## Conclusions

The breast cancer storytelling event provided an ideal opportunity for patients with breast cancer to meet and interact with researcher at various career stages successfully, using storytelling as a medium to share experiences. In this report we share our experiences and recommendations when considering the design of a patient engagement event with researchers. Findings from this PPIE activity while beneficial to all participants, has provided an educational resource for undergraduate students on the patient lived experience and educational needs at the beginning of a breast cancer diagnosis.

## Data Availability

All data produced in the present study are available upon reasonable request to the authors.

## Acknowledgments

The authors are sincerely grateful for the contributions from all patient participants who bravely shared their stories. We thank staff at the NHS Lanarkshire breast clinic for enabling the recruitment of patients to the engagement event. We acknowledge support from the North Lanarkshire council in providing access to the Summerlee Museum as a venue for hosting all three workshops. The authors acknowledge funding from the Engineering and Physical Sciences Research Council University of Strathclyde Impact Acceleration Account and the multiscale metrology suite (EP/V028960/1) for next-generation health nanotechnologies.

## Supplementary information

**Figure S 1.**
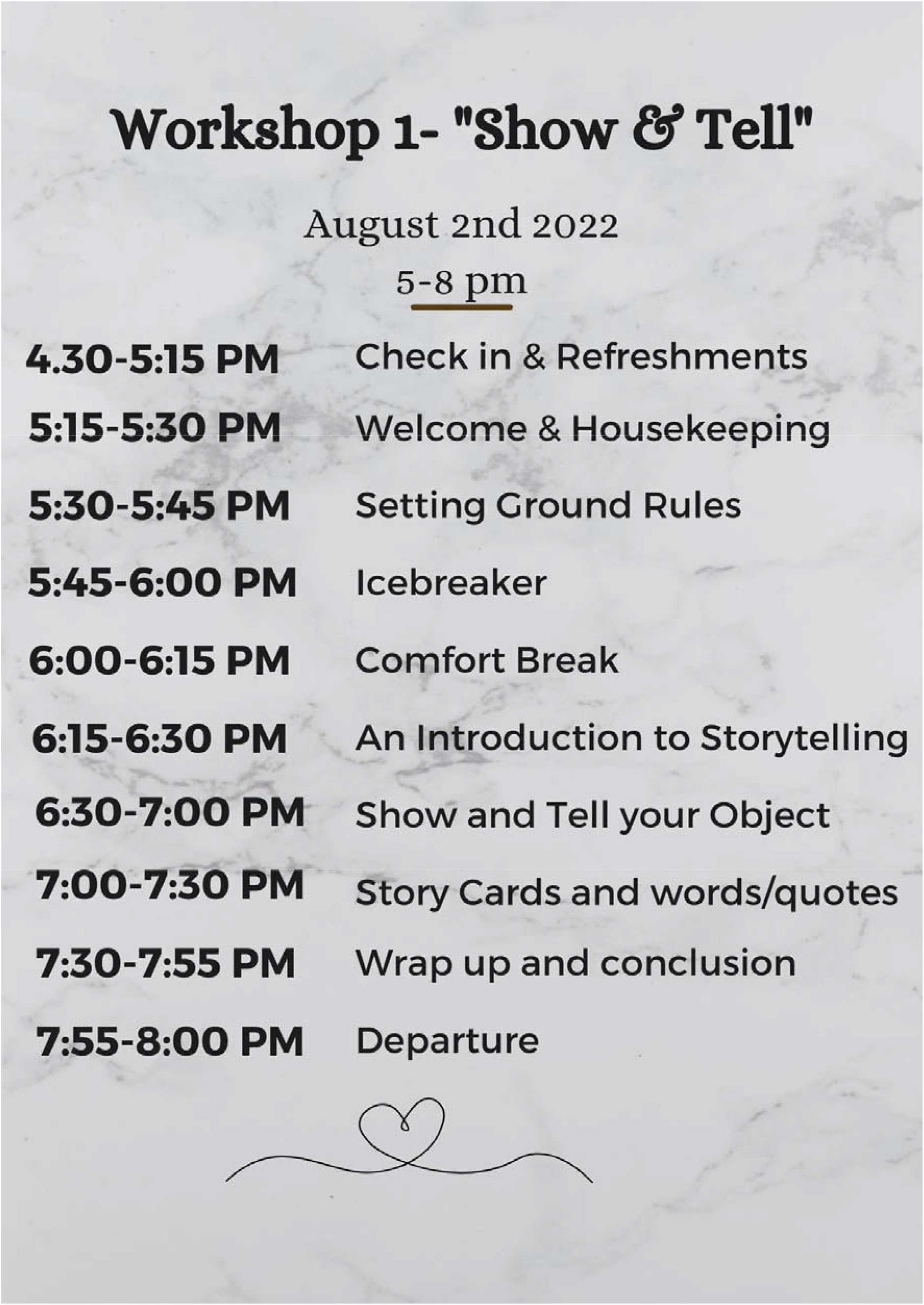
Workshop 1 running order provided to all participants on the day of the workshop

**Figure S 2.**
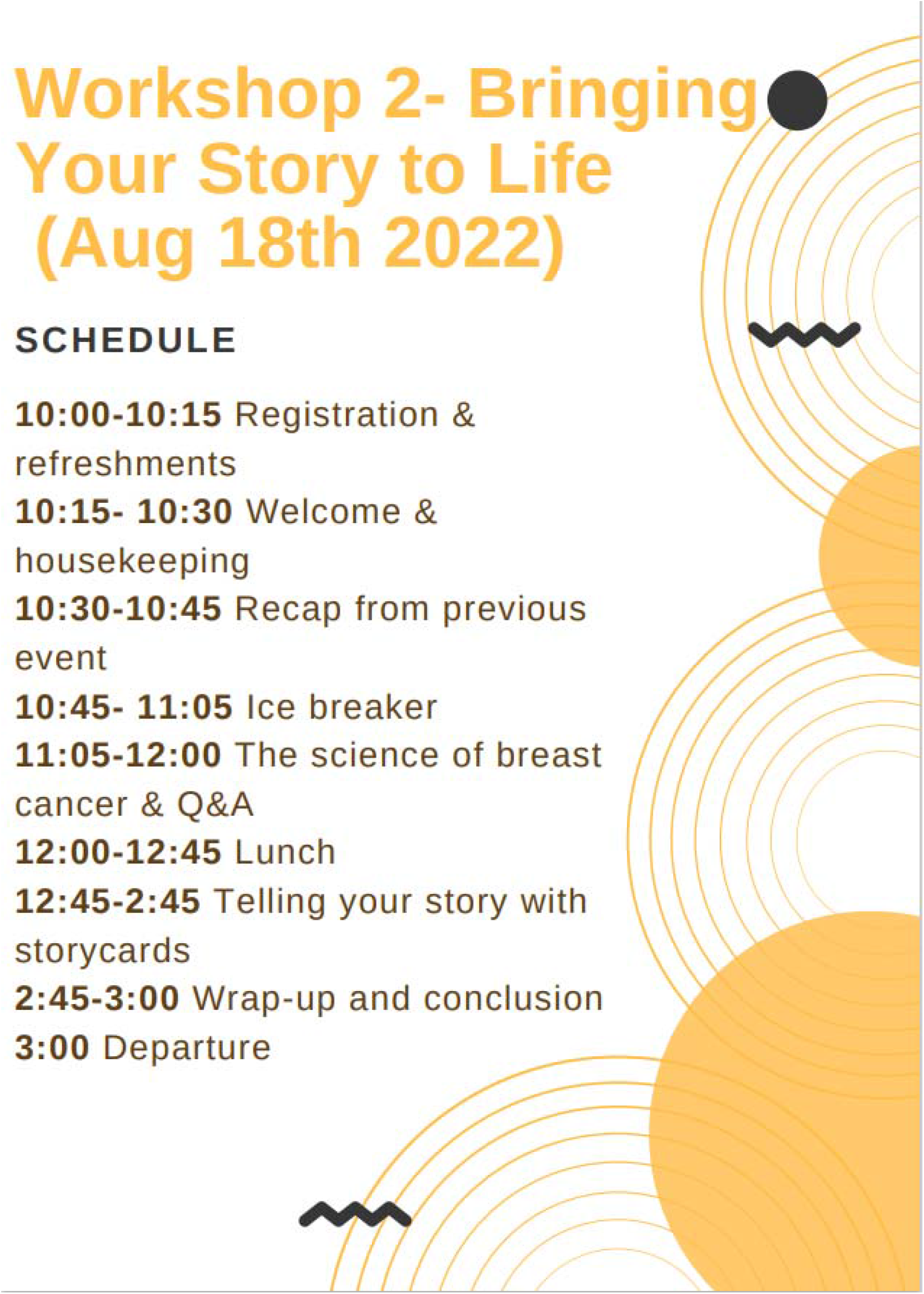
Workshop 2 running order provided to all participants on the day of the workshop

**Figure S 3.**
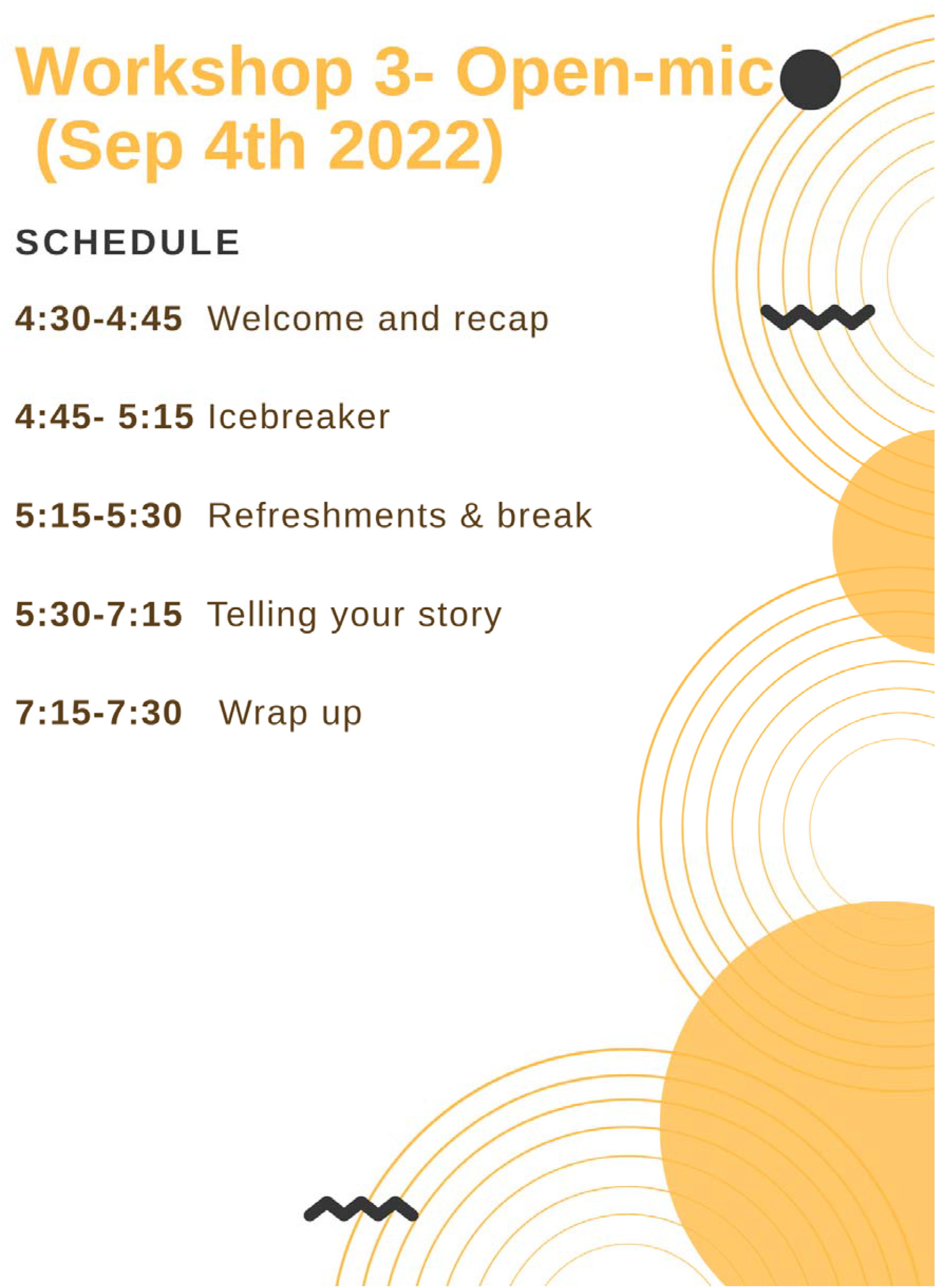
Workshop 3 running order provided to all participants on the day of the workshop

